# Quantifying the Tradeoff Between Energy Consumption and the Risk of Airborne Disease Transmission for Building HVAC Systems

**DOI:** 10.1101/2021.06.21.21259287

**Authors:** Michael J. Risbeck, Martin Z. Bazant, Zhanhong Jiang, Young M. Lee, Kirk H. Drees, Jonathan D. Douglas

## Abstract

It has been established that combinations of increased ventilation, improved filtration, and other HVAC techniques can reduce the likelihood of airborne disease transmission in buildings. However, with only qualitative guidance, it is difficult for building managers to make informed decisions. Furthermore, the possible actions almost always require additional energy consumption, which is generally not well characterized. To address this knowledge gap, we propose simplified physics-based models that can be used to quantify both the expected transmission rate and the associated energy consumption that result from HVAC system operation. By formulating all disinfection mechanisms in terms of “equivalent outdoor air”, a common basis is established for comparing and combining different strategies. The transmission rate can thus be modeled by considering the airborne concentration of infectious particles that would result from an infector in the space. Energy consumption is then estimated by considering the change in HVAC variables and applying standard analysis. To illustrate the insights provided by these models, we present examples of how the proposed analysis can be applied to specific spaces, highlighting the fact that underlying transmission risk and energy-optimal disinfection strategies can vary significantly based on space characteristics.

## 1. Introduction

With the recognition that COVID-19 and other diseases may be primarily transmitted through respiratory aerosols (Greenhalgh et al., 2021; Morawska and Milton, 2020; Bazant and Bush, 2021), it is crucial to understand how buildings and HVAC systems can influence the risk of indoor airborne transmission (Morawska et al., 2021). While it is tempting from a practical standpoint to impose one-size-fits-all limits on occupancy or social distance, the unfortunate reality is that indoor air conditions vary significantly with climate, building type, and use. Thus, no single recommendation can be universally applicable. Nevertheless, we show that useful decision-making guidelines can be derived from simple mathematical models of indoor disease transmission and HVAC operation. Our analysis shows the importance of considering *both* the risk of transmission *and* the impact on energy consumption associated with possible courses of action.

### 1.1. Airborne Disease Transmission

When individuals speak or exhale, they release many particles into the air of varying sizes (Wells and Wells, 1936; Duguid, 1946), depending both on mode of respiration (Morawska et al., 2009; Johnson et al., 2011) and the volume of vocalization (Asadi et al., 2019, 2020). These particles consist of liquid droplets (partially dried “droplet nuclei”) resulting from fragmentation and evaporation of respiratory fluids (mixtures of water, proteins, and salts), such as airway mucus (Johnson and Morawska, 2009) and saliva (Abkarian et al., 2020). In individuals who are actively infected with a respiratory disease, these droplets also contain viruses or bacteria capable of transmitting the infection (Buonanno et al., 2020). Large droplets (up to mm in diameter), which are most apparent in coughs and sneezes (Wells et al., 1939), tend to quickly settle to the ground from respiratory plumes (Bourouiba et al., 2014; Rosti et al., 2020). However, smaller aerosol particles (between 0.1 and 10 µm in diameter), which have much higher (number) concentrations in exhaled air, can remain suspended in the air for long periods, exceeding typical ventilation air change times. In indoor environments, these particles are spread throughout a space due to natural air currents and recirculation provided by the HVAC system (Bhagat et al., 2020; Linden, 1999; Foster and Kinzel, 2021), thus providing a potential route for disease transmission to susceptible individuals in the space who inhale them and are exposed to the pathogens they contain. Although not all diseases can be spread in this way, the airborne transmission pathway has been identified for many especially contagious respiratory diseases (tuberculosis, measles, influenza, SARS-CoV), and it has increasingly been recognized as the primary transmission route for SARS-CoV-2, the virus responsible for the COVID-19 pandemic (Bazant and Bush, 2021; Greenhalgh et al., 2021; Morawska and Milton, 2020).

Given the significance of indoor airborne transmission, it is of great interest for public health to reduce this risk as much as possible. An obvious way to reduce the magnitude of airborne transmission in buildings is to restrict the number of occupants in the building. For example, reducing occupancy by a given percentage provides a corresponding relative reduction in infection risk, but ultimately risk should be assessed on a case-by-case basis, as one building operating at 100% occupancy may be just as safe as another building at 10% occupancy. Screening of temperature or other symptoms at building entrances can significantly reduce the probability that an infectious individual enters the space, but such methods are clearly inadequate, especially given the prevalence of asymptomatic or pre-symptomatic transmission of COVID-19 (Gandhi et al., 2020; Abkarian et al., 2020). Requiring occupants to wear face masks provides significant reduction in airborne transmission risk by adsorbing the momentum of exhaled air and filtering out a significant fraction of potentially infectious aerosols, both when an infector exhales and again when a susceptible inhales (Bazant and Bush, 2021; Konda et al., 2020).

On the other hand, some strategies that have been employed during the COVID-19 pandemic have had very little impact on aerosol-based transmission. For example, enforcing six-foot social distancing or installing plexiglass dividers may help mitigate short-range airborne transmission in respiratory jets (Abkarian et al., 2020; Yang et al., 2020; Bazant and Bush, 2021) or large drop transmission in coughs and sneezes (Bourouiba et al., 2014; Rosti et al., 2020), but unless these measures significantly restrict the ability of air to flow from infectors to susceptibles, they have little effect on long-range airborne transmission. The importance of mitigating *airborne* transmission in buildings is underscored by recent statistical analyses of COVID-19 transmission in schools: although there was no significant effect of 3-foot versus 6-foot distancing (van den Berg et al., 2021) or barriers between desks, reduced transmission was observed as a result of mask use and ventilation improvements, especially if coupled with improved air filtration (Gettings et al., 2021). These observations suggest that longer-range aerosol-based airborne transmission is more significant than other shorter-range transmission routes, thus highlighting the importance of designing and operating HVAC systems not only for comfort and energy efficiency, but also for reduced risk of airborne disease transmission.

### 1.2. HVAC Mitigation Strategies

Beyond operational restrictions, building HVAC systems can be operated so as to reduce the concentration of infectious aerosols in the air. Through ventilation, filtration, and sedimentation (Bazant and Bush, 2021), some aerosols are simply removed from the indoor air, along with any infectious material they contain. Alternatively, ultraviolet radiation (García de Abajo et al., 2020; Heßling et al., 2020) can be used to deactivate pathogens within the particles, in addition to natural processes, such as exposure to concentrated solutes (Lin and Marr, 2019). Although these particles remain in the air, they no longer pose an infection risk. Finally, standalone equipment employing any of these mechanisms can be installed in the space to provide additional mitigation beyond what is available from the HVAC system. The main challenge to HVAC practitioners is thus in understanding how to assess the level of risk in a space and how to decide what additional measures to take given the risk reduction associated with each strategy.

An additional complication is that essentially all HVAC measures that reduce indoor airborne infection risk also require consumption of additional energy. Though increased energy use may be tolerable during rare pandemic events, assessing the tradeoff between energy and infection risk is critical to guiding decision-making in the context of long-term public health and sustainability. Furthermore, the presence of seasonal endemics (primarily influenza) means that buildings will need the flexibility to switch between strategies focusing on infection risk reduction or energy efficiency on a regular basis. To this end, some studies in the past have examined the energy impact of various HVAC infection control measures. For example, Azimi and Stephens (2013) compared the energy cost of extra ventilation to that of higher-efficiency filtration and concluded that filtration is more cost-effective for most climates. Other studies (Orme, 2001; Santos and Leal, 2012; MacNaughton et al., 2015) have assessed the cost of extra ventilation (although in the context of general indoor air quality rather than infection control in particular). However, the majority of past work has examined either indoor infection mitigation or building energy consumption in isolation, and so it is important to unify this analysis to account for the tight coupling between the two effects.

### 1.3. Overview of the Paper

Based on the previous discussion, it is clear that building managers and other stakeholders need to take actions to mitigate airborne transmission in buildings while balancing associated energy and other costs. Indeed, ASHRAE has recommended that building managers “select control options, including standalone filters and air cleaners, that provide desired exposure reduction while minimizing associated energy penalties” (ASHRAE Epidemic Task Force, 2021b). The overall goal of this paper is to provide a straightforward quantitative means to perform exactly that assessment and to illustrate potential conclusions that can be drawn from it.

The remainder of this paper is structured as follows. In Section 2, we present background and mathematical modeling to assess infection risk and energy consumption. This analysis includes models for the impact of changing HVAC control and operational variables on infection risk and energy consumption. Throughout the discussion, we provide appropriate units of measure for each model variable (but of course dimensionally equivalent units can be substituted as appropriate). In Section 3 we then apply the models from the previous section to various example cases and examine key trends. Finally, we conclude and discuss future extensions in Section 4.

## 2. Modeling and Analysis

Although disease transmission and infection is a complicated and inherently stochastic process, past work in the literature has shown that average infection rates and other related quantities can be modeled and predicted. Similarly, detailed analysis of building energy consumption is a very broad topic, but by focusing only on the components of the HVAC system that change in response to infection control measures, changes in energy consumption can be quantitatively modeled. In the interest of brevity, we focus on steady-state analysis for this paper, but dynamic extensions are straightforward (albeit more cumbersome). We start with a brief background and then present the models in forms tailored to our current use case.

### 2.1. Modeling Background

Mathematical modeling of airborne disease transmission dates back to the work of Wells (1955) and Riley et al. (1978). The basic premise of the Wells-Riley model and its various extensions (Gammaitoni and Nucci, 1997; Beggs et al., 2003; Nicas et al., 2005; Noakes and Sleigh, 2009; Stilianakis and Drossinos, 2010; Bazant and Bush, 2021) is to estimate the average concentration of infectious particles in the air, calculate a total dose of infectious particles received by each susceptible individual, and then determine the probability that the individual becomes infected. The infectious particle concentration is generally obtained by assuming the air in a space is well-mixed and solving for the concentration from a material balance. The dose received by an individual is then calculated from this concentration and can be expressed in any appropriate amount unit of pathogen content, e.g. the number of virions or bacteria, or the total mass of infectious material. Following Wells (1955), it is customary to express these concentrations in terms of the “infection quantum”, which captures the average amount of pathogen required to cause an infection. Infectious dose is then expressed in units of “quanta”, i.e., multiples of the quantum. In the context of the Wells-Riley model (which is valid for exposure times shorter than the mean incubation time of the disease), the infection quantum may be more precisely interpreted as the amount of pathogen carrying an infection probability of 1 − exp(−1) ≈ 63%, based on a simple exponential relaxation to steady state (Noakes and Sleigh, 2009).

Mathematically, these relationships are expressed as

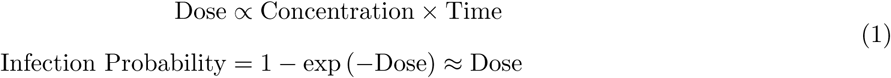

In the first equation, the constant of proportionality depends primarily on the individual’s breathing rate, but it can also include other factors such as the filtration provided by a mask. In the second equation, the exponential dependence reflects the fact that infection becomes increasingly certain (i.e., approaches 100%) as the dose increases. The linear approximation (which is accurate within 10% relative error for doses less than 0.2 quanta), can be used to derive a simple safety guideline to limit airborne transmission in any indoor space (Bazant and Bush, 2021), and we use a similar approach here to quantify overall infection risk in each space. This probabilistic formulation ultimately gives a distribution for the number of additional infections across all building occupants, with the average value calculated as the sum of each individual’s probability.

The well-mixed assumption is widely used in HVAC system design to describe mass and energy balances in buildings, and, aside from some corrections for short-range respiratory flows (Abkarian et al., 2020; Yang et al., 2020) and fluctuations in indoor convection (Bhagat et al., 2020), it can also be justified theoretically (Bazant and Bush, 2021) and through simulations (Foster and Kinzel, 2021) as a useful first approximation in modeling airborne disease transmission. Recent extensions to this basic premise include the consideration of droplet size distributions depending on respiratory activity (Bazant and Bush, 2021), inclusion additional removal sources (Azimi and Stephens, 2013; Dai and Zhao, 2020; Shen et al., 2021), considering ventilation effectiveness and social distance (Sun and Zhai, 2020), accounting for the transient dynamics associated with the infectious particle concentration (Stilianakis and Drossinos, 2010), using CO_2_ concentration measurements to infer occupancy or exhaled aerosol concentrations (Rudnick and Milton, 2003; Peng and Jimenez, 2021), and simulating partitioning and recirculation effects (Pease et al., 2021). For our part, we take a similar approach but introduce a more systematic treatment of the HVAC removal sources that allows us to also account for corresponding energy consumption incurred by various mitigation strategies for airborne disease transmission.

### 2.2. Equivalent Outdoor Air

To assist our modeling discussion, we start by defining the concept of equivalent outdoor air (EOA). As presented by the ASHRAE Epidemic Task Force (2021a), the EOA delivered by a series of disinfection devices is the equivalent volume of outdoor air that, when circulated through the space, would remove the same amount of infectious particles as removed (or deactivated) by the device(s). This formulation allows for a direct comparison of the disinfection provided by various devices that may operate on different principles.

For most devices, the EOA can be characterized in terms of the total flow *f* (m^3^*/*h) through the device and a constant infectious-particle removal efficiency *η* expressed as a value between zero and one. The resulting EOA flow is then given as 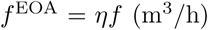. For example, in a ventilation system where 30% of of the air is vented to the outdoors and replaced with a corresponding volume of outdoor air, we have *η* = 0.3. Similarly, for filtration devices, *η* is the fraction of infectious particles trapped by the filter, which can be calculated from the filter’s MERV rating and the size distribution of infectious particles. Previous studies for influenza (Azimi and Stephens, 2013) have found that roughly 20% of viral material resides in the E1 size range (0.3 µm to 1 µm), 29% in the E2 size range (1 µm to 3 µm), and 51% in the E3 size range (3 µm to 10 µm). Using these values, the overall filtration efficiency is given as

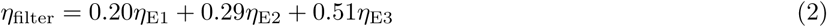

in which the *η*_∗_ values are the filtration efficiencies for each size range as defined in the MERV standard. Using those values gives total efficiencies of *η* = 0.56 for a MERV 8 filter and *η* = 0.90 for a MERV 13 filter. It should be noted that the infectiousness of entrained pathogens may vary with drop size and humidity (Bazant and Bush, 2021; Lin and Marr, 2019), depending on disinfection kinetics and the relative ease of penetrating the respiratory tract, but the values presented here based on viral load distributions for influenza can serve as a rough guide for the size-dependence of pathogen filtration efficiency in other scenarios. Indeed, most existing models of airborne transmission tend to neglect the size dependence of any pathogen removal processes, other than sedimentation (Stilianakis and Drossinos, 2010; Bazant and Bush, 2021). Alternatively, due to the inherent uncertainty in both filtration efficiency and particle size distribution, one could opt for a conservative bound by assuming the lowest efficiency for that filter type, i.e.,

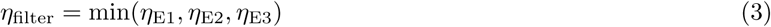

which would give a worst-case estimate of the EOA delivery for that filter (and thus a conservative bound on the resulting infection risk). For ultraviolet germicidal irradiation (UVGI) devices, proper configuration can give *η* ≈ 1 for the air flowing through the device at design conditions. However, if radiation intensity is not sufficiently high, then the dose response of the pathogen may need to be considered (Kowalski, 2010), which would result in *η* (and thus also *f* ^EOA^) being a nonlinear function of *f*.

The main caveat is that when disinfection devices operate in series, the EOA calculation must account for the fact that downstream devices cannot remove infectious particles that have already been removed upstream. Thus, the total efficiency calculation must account for this effect. As an example, in a typical AHU configuration, the return air stream is partially ventilated with a fraction *η*_vent_ and then the resulting mixed stream is passed through a filter with efficiency *η*_filter_. The overall efficiency *η*_AHU_ for the AHU is thus given by

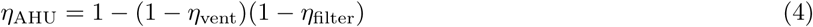

Were there any other devices in the AHU duct (e.g., UVGI lamps), their efficiencies would be included as additional 1 − *η*_∗_ terms in the product on the right-hand side. The resulting EOA flow is thus 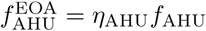.

Finally, we note that the natural deposition and deactivation of the infectious particles can be considered in the EOA framework. Assuming that the particles deposit by sedimentation at a rate of *β*_deposition_ (h^−1^) and the viruses within the particle decay at a rate *β*_deactivation_ (h^−1^), we have a corresponding EOA given by

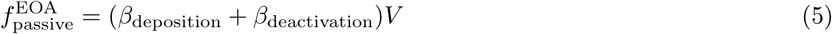

where *V* is the total volume of air in the space. Both effects (especially deposition) do exhibit size dependence, which can be addressed similarly to the filtration efficiency to determine effective values for the parameters based on the size distribution of interest. We refer to deposition and deactivation as “passive” disinfection, as they occur regardless of the operation of the HVAC system. In many spaces, for COVID-19, they account for a modest correction to the EOA rate on the order of 0.75 to 1.5 air changes per hour (ACH) with weak dependence on humidity (Yang and Marr, 2011; Lin and Marr, 2019; Bazant and Bush, 2021). Accounting for these processes in the EOA framework can often simplify analysis and helps to provide context for the disinfection efficacy of the HVAC system and other devices.

### 2.3. Infectious Particle Concentration

To assess airborne infection risk in a space, we start with a simple model for the concentration of infectious particles in the air. For this purpose, we assume that the space is well-mixed, i.e., that the concentration of infectious particles (expressed in units of quanta*/*m^3^) is uniform. Similar to any airborne contaminant (PM, CO_2_, VOCs, etc.), the average concentration of infectious particles in the air is determined by the balance of generation and removal. The generation rate depends on the number, breathing rate, and mask usage of infectious individuals in the space, while the removal rate is determined by a combination of passive physical processes (deposition and deactivation), operation of the HVAC system (filtration and ventilation), and any other disinfection technologies deployed within the space.

To describe this mathematically, we let *N*_*I*_ be the number of infectors (#) in the space, *C*_*I*_ be the net infectious-particle concentration in the infectors’ exhaled breath (quanta*/*m^3^), *η*_mask_ be the filtration efficiency of the infectors’ masks (0 to 1, with *η*_mask_ = 0 if not wearing masks), *f*_*b*_ be the average breathing flow rate (m^3^*/*h, exhaled or inhaled volume per time), and 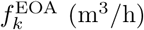 be the EOA flow to the zone from source *k*. Under steady-state conditions, a balance between generation and removal gives the simple relationship

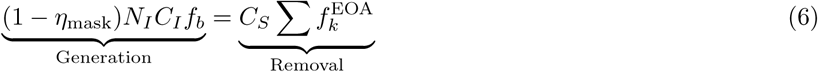

This equation can then be solved for *C*_*S*_, which is the average airborne infectious-particle concentration in the space (quanta*/*m^3^) and thus also the concentration in the susceptibles’ inhaled air. This value can then be used to calculate the expected transmission rate in a given space.

#### 2.3.1. Discussion

From Eq. (6), the usefulness of the EOA approach is apparent: by converting each disinfection source *k* to its equivalent outdoor air flow 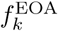, the resulting removal rate is simply the product of that flow with the infectious-particle concentration in the air. For most indoor spaces, there will be two primary disinfection sources *k* ∈ {passive, AHU} (recalling that passive effects include deposition and deactivation), while the AHU accounts for ventilation, in-duct filtration, and any other in-duct technologies. For spaces where extra disinfection is provided, e.g., by means of in-zone filtration devices, the impact on infectious particle concentration can be determined by adding an additional term to the sum. Note that when combining different filtration sources, there is potentially a shift in the resulting particle size distribution in the air, which means the resulting filtration efficiencies are slightly different from their original values. For typical cases where the supplemental sources are rated MERV13 or higher, this effect can generally be neglected due to the much weaker size-dependence of filtration efficiency for these types (ASHRAE Epidemic Task Force, 2021a). However, when there are multiple independent filtration sources rated MERV8 or lower, the resulting particle concentrations may need to be considered on a size-resolved basis.

#### 2.3.2. Limitations

Before moving on, we must briefly comment on the validity of the well-mixed assumption used to derive Eq. (6). Because we are modeling exhaled particulates, additional justification is needed to assume perfect mixing compared to, e.g., modeling gaseous species. In still air, the movement of such particles is due primarily to diffusion and buoyancy-driven natural convection, which would suggest formation of a high-concentration plume in the near vicinity of each source based on the relevant timescales. However, when the HVAC system is active, the forced circulation of air leads to much faster particle transport and rapid mixing throughout the space by typically turbulent flows. Furthermore, because exhaled air is warmer than typical indoor air, buoyancy leads to natural convection transporting exhaled particles upwards around an infected person (Bhagat et al., 2020), thus giving them additional time to mix with the air in the space before gradually moving back down to the breathing zone where they may be inhaled by susceptibles. Given these effects, theoretical arguments (Bazant and Bush, 2021), fluid-dynamical simulations (Foster and Kinzel, 2021) and experimental evidence have all shown that beyond roughly one meter of distance from an infector, the well-mixed assumption is typically a very good approximation and can at least serve as a starting point for analysis of local or other nonuniform effects. Thus, we consider the well-mixed assumption to be valid for our use case.

### 2.4. Transmission Rate Calculation

Our ultimate goal in infection-risk modeling is to determine the number or rate of new transmissions that occur in a building. As discussed in Section 2.1, the general approach would be to calculate each individual’s infectious dose and sum the infection probability across all individuals in the building to determine the average number of transmissions. Unfortunately, this individual-centric analysis is difficult because it requires knowledge of each individual’s location throughout the day to track their specific exposure. However, by making use of the linear approximation to the exponential probability relationship, we can instead adopt a *space-centric* approach in which we calculate the total dose received by individuals in that space and then sum over all spaces in the building. In addition to simplifying analysis, this strategy is more helpful to building managers, as it allows them to identify which specific spaces are in need of additional disinfection.

Mathematically, the total infectious dose *D*_*S*_ (quanta) received across all susceptible occupants in a space is given by

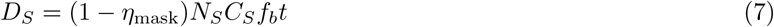

in which *N*_*S*_ are the number of susceptibles in the space (#), *t* is the average amount of time each susceptible spends in the space (h), and *C*_*S*_ is the infectious particle concentration in the space (quanta*/*m^3^). The remaining parameters *η*_mask_ and *f*_*b*_ are the mask filtration efficiency and breathing rate as before. Consistent with Eq. (1) and the definition of the infectious quantum, each quantum of total dose is expected to cause one new transmission to the susceptible population of the space. Thus, the expected number of transmissions *N*_*T*_ (#, but can be fractional) is given by

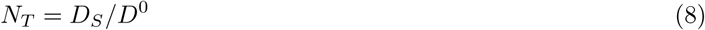

in which *D*^0^ = 1 quanta/transmission is the appropriate scaling factor. Finally, we can then combine Eqs. (6) to (8) and arrive at the model

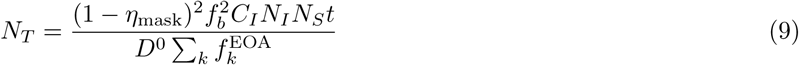

We note that this value is space-centric in that it considers only the transmissions that occur due to occupant exposure while they are in the specific space.

To compare underlying infection risk across spaces, it is sometimes useful to consider the *indoor reproductive number* associated with the disease transmission (Bazant and Bush, 2021), which is given by *R* = *N*_*T*_ */N*_*I*_. In this formulation, *R* (-) thus gives the average number of additional infections caused by each infector in the space. However, when comparing spaces with vastly different occupancy it is often more useful to work with *N*_*T*_ directly, as it accounts for the number of occupants and is thus a more direct measure of overall infection risk.

#### 2.4.1. Discussion

In Eq. (9), we can generally split the variables into three groups: basic properties of the disease and humans (*C*_*I*_, *f*_*b*_, and *D*^0^); policy decisions made by building owners (*η*_mask_, *N*_*I*_, *N*_*S*_, and *t*); and, HVAC operational variables (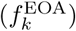). In the first group, we note that the infectors’ exhaled infectious particle concentration *C*_*I*_ can vary over multiple orders of magnitude depending on activity of the infectors, ranging from roughly 10 quanta*/*m^3^ for quiet breathing to 100 quanta*/*m^3^ for speaking, and even as high as 1000 quanta*/*m^3^ for singing (Bazant and Bush, 2021). This strong dependence is due to the physical processes that generate the respiratory droplets. By contrast, the breathing rate *f*_*b*_ is generally in the range of 0.5 to 1 m^3^*/*h for light activity, although it can be as high as 3 m^3^*/*h during vigorous exercise (Agency, 2011). We do note that the infection rate depends on the *square* of the breathing rate, as it is relevant for both generation by infectors and exposure to susceptibles. As mentioned previously, the scaling parameter *D*^0^ is generally set equal to 1 transmission/quanta, but it could potentially be used to account for different infectivity of variants instead of adjusting *C*_*I*_.

For the policy parameters, it is important to recognize that the mask filtration efficiency *η*_mask_ should account not only for the physical filtration properties of the material but also the fit of the mask as worn by individuals. Thus, although a well-fitting mask can remove upwards of 95% of exhaled particles, the ability of air to escape around the edges of a loose fitting mask can reduce the effective efficiency down to between 50% and 90% (Bazant and Bush, 2021). Fortunately, because of the quadratic dependence on 1 − *η*_mask_, even a modest *η*_mask_ = 0.7 reduces infection risk by a factor of 1*/*(1 − 0.7)^2^ ≈ 10. Thus, by requiring occupants to wear masks, building managers can allow higher occupancy levels (*N*_*I*_ and *N*_*S*_) or durations (*t*) while still maintaining satisfactory infection risk. The remaining variables 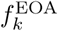 are related to the operation of the HVAC system and will be discussed in more detail in Section 2.5.

Finally, we note that the calculation of *N*_*T*_ in Eq. (8) as the expected number of transmissions in the space derives from the linear approximation in Eq. (1). To illustrate, we assume that there are *N*_*S*_ total susceptible individuals (each indexed by *i*) that are exposed to *individual* doses *D*_*i*_ (quanta) within the space of interest after having already been received doses 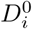 (quanta) prior to entering the space. Thus, each individual’s prior infection probability 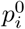 and final infection probability *p*_*i*_ (0 to 1) are given by

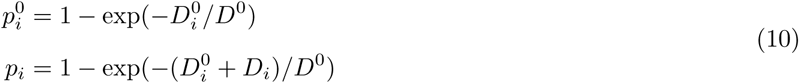

We can then calculate the expected number of transmissions in the space as 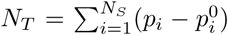. Unfortunately, this expression is not particularly useful because we have to know each individual’s prior exposure 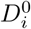. However, if we assume that each individual dose is sufficiently small 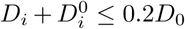, we can approximate as follows:

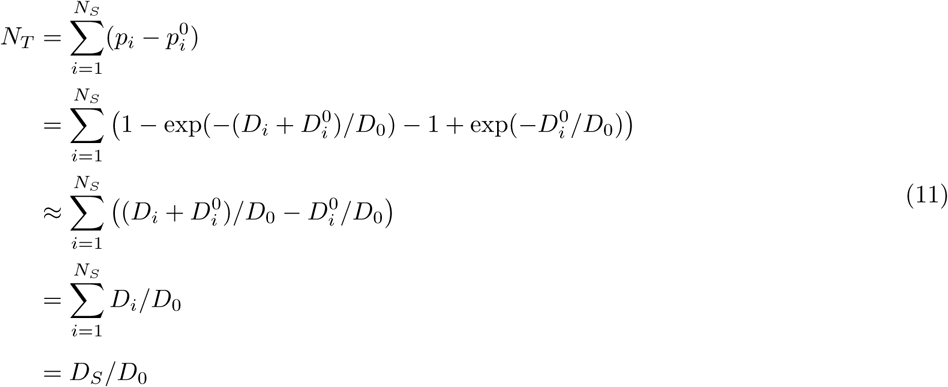

which gives *N*_*T*_ as in Eq. (8). The important benefit is that the individuals’ prior exposures have canceled out, and thus we can focus only on what occurs within the spaces being considered. Over the daily time periods relevant to building occupants, we expect that this linear approximation is highly accurate, as higher individual doses would indicate an unacceptably high transmission rate. However, we note also that the third line of Eq. (11) is a strict *overapproximation* (due to concavity of the exponential function), and thus under all circumstances, *N*_*T*_ ≤ *D*_*S*_*/D*_0_. Indeed, using this conservative approximation is appropriate in our context, as the overall goal is to maintain total infectious doses as small as possible.

#### 2.4.2. Limitations

To close this section, we briefly discuss some limitations of Eq. (9) and how they could potentially be accounted for. First, because the equation assumes steady state conditions in the space, it inherently ignores transient dynamic effects. For example, when an infector first enters the space, the infectious particle concentration *C*_*S*_ exponentially approaches the steady-state concentration with a time constant 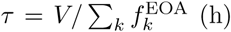; when they leave the space, there is a symmetric exponential relaxation back down to zero. Thus, if the infector only spends a small amount of time in the space (e.g., as might be typical in retail spaces), then the infectious particle concentration will never reach its steady-state value, which means Eq. (9) will overestimate the resulting infection rate. However, if the infector spends at least 3*τ* time in the space, then the steady-state approximation will be highly accurate due to the dynamic symmetry. To account for this fact, the variable *t* in Eq. (9) should be interpreted as the average amount of time each *infector* spends in the space. Where additional time-varying information is available, a more detailed dynamic model can be simulated, but in most cases the steady-state analysis will be sufficient. Furthermore, to account for variables that change on a slower timescale, the total time interval can be split into multiple separate periods, and the steady-state analysis can be applied separately to each period.

Second, by making the well-mixed assumption, the model Eq. (9) inherently neglects the extra risk associated with short-range transmissions, specifically when susceptibles directly inhale the exhaled breath of infectors. When masks are not worn, the exhaled aerosols are transported by turbulent respiratory jets, whose momentum and droplet density are amplified by speaking or singing (Abkarian et al., 2020). In such short-range aerosol transmissions, the infectious particle concentration received by the susceptible occupant may be closer to the infector’s full exhaled concentration *C*_*I*_ than the average concentration *C*_*S*_ in the space. Social distancing provides some protection against the enhanced risk of short-range transmission via respiratory jets, as their infectiousness scales inversely with distance (Yang et al., 2020). In contrast, when masks are worn, exhaled breath loses most of its momentum by passing either through or around the filtration material, and the warm exhaled air tends to quickly rise by natural convection and become mixed throughout the space (Bhagat et al., 2020). The excess risk from these respiratory flows can be modeled with a correction to Eq. (9), which accounts for the relative importance of short-range versus long-range aerosol transmission, as discussed by Bazant and Bush (2021).

Third, when Eq. (9) is applied to the full space served by a single AHU, the well-mixed assumption fails to account for the fact that the space may be partitioned into multiple rooms that are largely isolated other than shared supply and return airstreams. As discussed in Pease et al. (2021), models that account for partitioning predict a higher infectious particle concentration in rooms with an active infector and lower concentrations in rooms without an infector as compared to the uniform model in Eq. (9). However, it can be shown that if the per-room airflow and occupancy variables (*N*_*I*_, *N*_*S*_, and *f* ^EOA^) are all proportional to each room’s floor area, then the total infectious dose *D*_*S*_ calculated by the more detailed partitioned model is the same as calculated by Eq. (9). Therefore, it is only necessary to account for partitioning when there is significant heterogeneity in airflow or occupant density within the individual rooms.

Finally, we note that although the predicted transmission and reproductive numbers *N*_*T*_ and *R* are useful for assessing relative infection risk, they are inherently less value for absolute interpretation. Specifically, it is challenging to define a fixed threshold for either variable below which the space is considered “safe”. In theory, maintenance of *R <* 1 causes the disease die out and prevent runaway spread, which would suggest that value as a safety target. However, because the computed *R* value considers only time spent in the space of interest, *R* = 1 often corresponds to unacceptably high infection risk, since occupants could receive additional exposure in other spaces or buildings. Furthermore, due to the significant uncertainty in the exhaled quanta concentration *C*_*I*_, the estimated values of *R* and *N*_*T*_ could be incorrect by an order of magnitude or more, and thus any target would require a significant margin for error. In general, we regard *N*_*T*_ as the best direct metric for comparing infection risk across spaces, because it inherently accounts for different levels of occupancy in the spaces being considered. From this standpoint, mitigation efforts should be prioritized in terms of how much they reduce total expected transmissions (e.g., as illustrated in Section 3.3). Unfortunately, an absolute interpretation of *N*_*T*_ is even more challenging because the true value of *N*_*I*_ depends on many external factors including local community prevalence of infection and immunity, as well as entrance screening procedures that may prevent infected individuals from entering the space. Thus, rather than setting any specific absolute target for *N*_*T*_, the overall goal should be to maintain it as low as possible within the constraints imposed by other considerations (e.g., energy consumption, occupant comfort, and equipment capacities).

### 2.5. HVAC Measures and Energy Consumption

As discussed in the previous section, Eq. (9) provides a means to quantify the airborne infection risk in buildings. For a given set of disease and operational policy variables, the resulting infection rate in each space is inversely proportional to the total EOA delivered to this space. Thus, by choosing to operate their HVAC systems in a way that increases the flow of EOA, infection risk is thus reduced. Unfortunately, in almost all circumstances, such actions also increase energy consumption, and so it is important to choose the most energy-efficient disinfection strategies. In the following, we briefly discuss possible ways to increase EOA along with simple models to estimate the associated extra energy consumption. In all cases, the effect on indoor infection risk can be assessed by adjusting the values of 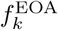 in the denominator of Eq. (9) in accordance with the change in EOA. Note, however, that these models do not account for equipment capacities or other limitations, and so it is always necessary to check that any possible modification does not adversely affect HVAC system operation or occupant comfort.

#### 2.5.1. Improved Filtration

One of the simplest ways to increase EOA delivery to a space is to install a filter with a higher rating. Indeed, the ASHRAE Epidemic Task Force (2021a) has recommended that filters be upgraded from the standard MERV 6 or MERV 8 to an efficiency of MERV 13 or higher. As discussed in Section 2.2, the primary effect of this change is to increase the filtration efficiency *η*_filter_, which in turn increases the EOA flow 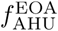 from the AHU. However, to calculate the increase in EOA, it is important to consider the series effects for ventilation (and any other AHU disinfection sources) as discussed in Section 2.2. The net result is that when the outdoor-air fraction is nonzero (which should always be the case during occupied hours), a filter upgrade will not provide the full increase in EOA, as the air stream has already been partially pre-cleaned before passing through the filter.

The primary energy effect of switching to a higher-efficiency filter is an increase in fan power due to the increased pressure rise needed to move air through the more efficient filter. This amount can be estimated via the design pressure drop across each filter type and the design pressure rise of the fan. Mathematically, we have

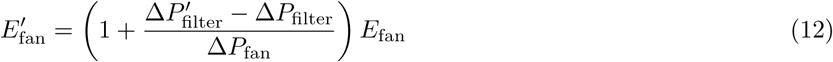

in which *E*_fan_ is the baseline fan energy consumption (kWh, which can be estimated from design flow and power values for the fan), Δ*P*_filter_ is the design pressure drop (Pa) of the original filter, 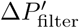 is the design pressure drop (Pa) of the new filter, Δ*P*_fan_ is the total design pressure rise (Pa) for the fan, and 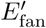 is the new energy consumption (kWh) of the fan after installing the new filter. This approach of assuming linear dependence of energy on pressure drop has been adopted in past studies (Montgomery et al., 2012; Azimi and Stephens, 2013) and is appropriate for variable-volume systems. However, other studies have shown that, particularly for smaller or constant-volume systems, changing the filter type can have a direct impact on total airflow, and thus both the EOA and energy implications can be difficult to predict (Stephens et al., 2010; Zaatari et al., 2014). In such cases, additional modeling may be necessary.

#### 2.5.2. Increased Ventilation

An alternative to improved filtration is increased ventilation, which delivers more clean outdoor air to the space and thus vents a corresponding amount of indoor air (which contains infectious particles when infectors are present). We note that throughout this paper, we use “ventilation” to refer specifically to *outdoor-air* ventilation, which thus does not include the airflow associated with recirculation. Just as in the case of improved filtration, it is important to account for the coupling between the two effects. Specifically, if the increased ventilation is provided only by increasing the *fraction* of outdoor air (i.e., increasing *η*_vent_ but keeping *f*_AHU_ constant), then the resulting effect on EOA may be negligible. This behavior will be illustrated in Section 3.1.

Similar to the delivered EOA, the energy consumption associated with increased ventilation can vary from negligible in mild outdoor conditions to very high in extreme conditions. Furthermore, the analysis is somewhat complicated by the fact that the increased energy consumption may be due to heating or cooling depending on the season. In general, the coil thermal load is calculated as the enthalpy difference between the supply and mixed air streams, with the “mixed” stream being the mixture of recirculating indoor and ventilation outdoor air (in which *η*_vent_ gives the fraction of outdoor air). Mathematically, the resulting energy consumption *E*_AHU_ (kWh) is given by

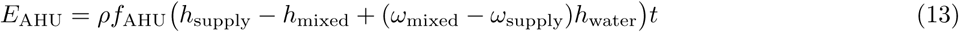

Here, the *h*_∗_ and *ω*_∗_ variables are the wet-air enthalpy (kJ*/*kg) and absolute humidity ratios (kg*/*kg) of the given air streams (which can be calculated from the corresponding temperatures *T*_∗_ (^◦^C) and humidities *φ*_∗_ (%) using psychrometric charts), while *ρ* is the standard density of air (kg*/*m^3^), *f*_AHU_ is the AHU supply flow (m^3^*/*h) as before, *h*_water_ is the enthalpy (kJ*/*kg) of *liquid* water at supply conditions, and *t* is the operating time (h). Note that a positive value of *E*_AHU_ indicates a heating load, while a negative value indicates cooling. When aggregating with other sources of energy consumption, this value should be appropriately scaled by the (inverse) coefficient of performance for cooling or efficiency for heating to convert to electricity equivalents.

To determine conditions of the supply air stream, *T*_supply_ can be set equal to its setpoint, while *φ*_supply_ is close to 100% during cooling conditions and the resulting mixture of *φ*_outdoor_ and *φ*_indoor_ during heating conditions. Slightly more complicated models are available to account for heat-transfer effects at the coils (Seem and House, 2010). In any case, to estimate the energy impact of additional ventilation, the given model evaluated for the different values of *η*_vent_ being considered. Indeed, accounting for the change in sensible and latent loads is important in the cooling season, especially in particularly moist climates where dehumidification is significant. We do note that for buildings employing energy-recovery ventilation, the analysis will need to be modified to account for the corresponding heat and mass transfer. Specifically, the mixed-air conditions should account for the temperature and humidity of the outdoor-air stream *after* it has passed through the energy-recovery device (which requires an appropriate heat-transfer model). In many cases, these devices can pre-condition the outdoor air, bringing it thermodynamically close to the recirculating air stream, and thus the resulting change in energy consumption from the baseline case can be essentially negligible (MacNaughton et al., 2015). On the other hand, buildings with these types of devices may already be operating at maximum ventilation rates, and thus this variable would not be adjusted for infection control.

#### 2.5.3. Additional Recirculation

As an alternative to improved filtration or increased ventilation, one strategy to deliver additional EOA to the space is simply to provide additional recirculating air. Because this air passes through the filter (and contains some fraction of outdoor air), it is partially cleaned. In this case, the filter efficiency *η*_filter_ stays the same, and the outdoor air fraction *η*_vent_ may stay the same or decrease depending on the particular control strategy, but the total flow rate *f*_AHU_ increases. Where equipment capacities permit, this approach can provide a much more dramatic change in EOA and often without significant energy penalty. Unfortunately, in many HVAC systems there may not be a direct mechanism to increase the recirculation rate. However, one case where this is possible is variable-volume systems in cooling mode: by raising the supply temperature setpoint, the zone temperature controllers will naturally increase airflow to the space. The precise amount of extra air will ultimately be determined by the temperature control loop, but it can be estimated as

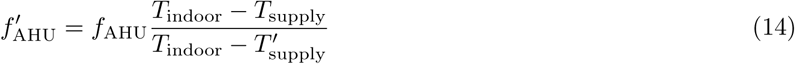

Here, 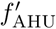 is the new airflow (m^3^*/*h), which is equal to the original airflow *f*_AHU_ (m^3^*/*h) multiplied by the ratio of temperature differences at the old and new supply temperatures *T*_supply_ and 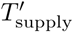 (both ^◦^C). This relationship is derived from the fact that, to maintain constant temperature in the zone, the net cooling load is constant. Thus, by equating the original cooling load (which is proportional to *f*_AHU_(*T*_indoor_ − *T*_supply_) to the new cooling load (which is proportional to 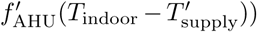, we can rearrange to find Eq. (14). Note that as the above equation implies, it is also possible to achieve the same effect by lowering the indoor zone temperature *T*_indoor_ (via its setpoint), but such a change may have an impact on occupant comfort. In any case, once the new AHU airflow has been calculated, the resulting EOA can be calculated as before by multiplying by the effective AHU removal efficiency *η*_AHU_.

The energy analysis associated with additional recirculation is the most complicated, as it affects both fan power and coil energy. For this purpose, we consider again the case where the increased flow is provided by increasing the supply temperature setpoint. For fan power, the standard model assumes a cubic dependence of energy consumption on flow. Thus, on a relative basis, we have

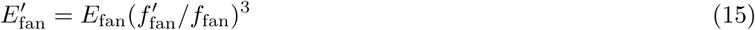

i.e., that the new energy consumption 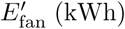 is equal to the original energy consumption *E*_fan_ (kWh) multiplied by the ratio of the new and old flows 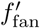 and *f*_fan_ (both m^3^*/*h) raised to the third power, with the new flow calculated as above in Eq. (14). To estimate the heating and cooling energy consumption, the best method is to simply apply Eq. (13) to the possible cases as when considering increased ventilation. The main difference is now that *T*_supply_ will be the primary variable being adjusted. We note that for this case, the fan and AHU coil energies will often move in opposite directions as supply temperature is increased, with fan power steadily increasing (due to increased flow) and coil power either staying the same or decreasing (due to the reduced dehumidification load). Thus, total energy may increase or slightly decrease depending on the magnitude of these effects. We show examples of this behavior in Section 3.2.

#### 2.5.4. In-Zone Disinfection

Finally, if it is not possible to provide sufficient EOA via the three previous HVAC mechanisms (e.g., due to equipment capacity limitations or operational concerns), an additional option is to simply install standalone disinfection devices within the zone. These devices operate on their own stream of air, and they generally operate via filtration, UV disinfecion, or a combination of the two. For example, an in-zone HEPA filtration unit (*η*_filter_ = 0.999) with an airflow of *f*_in-zone_ = 1, 000 cfm would deliver an EOA of 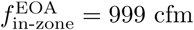. Fortunately, the energy analysis for these devices is the most straightforward, as the baseline consumption is zero and the new consumption is simply the rated power of the device multiplied by the time period which it is active. Since these devices can be expensive, it is important to identify the space where they will have the most impact, which can be determined by the space for which inclusion of the device provides the greatest reduction in expected transmissions as predicted by Eq. (9). Such analysis is illustrated in Section 3.3.

## 3. Case Studies

To demonstrate the utility of the proposed modeling approach, we present a set of example case studies that illustrate the potential insights that can be gained from the models. These calculations are not intended as general conclusions that would apply to all spaces or buildings of a given type but rather to highlight that the analysis is straightforward to apply and should be applied separately for each space. Indeed, the example cases show that there is significant variation in infection risk and energy consumption across different buildings and climates, which further underscores the fact that one-size-fits-all guidelines are not sufficient.

## 3.1. Infection Risk

As a first example case, we show how the number of transmissions varies with EOA delivery for different space types. To standardize results, we assume 100 total occupants with one infector and no one wearing masks. Space dimensions are then calculated from representative occupant densities assuming 10 ft ceilings. We use a breathing rate of 0.75 m^3^*/*h (which corresponds to light activity) and quanta generation rates are chosen based on typical levels of talking. Key parameters are shown in Table 1. Using Eq. (9), the expected number of transmissions *N*_*T*_ in each space can be calculated as a function of total EOA delivery, assuming each space is occupied for 8 hours each day. (Because we assume one infector, the displayed values of *N*_*T*_ also correspond to the daily indoor reproductive number *R*). To estimate uncertainty, we assume that the exhaled quanta concentration may be higher or lower by a factor of 3 compared to the baseline value in Table 1. These results are shown in Fig. 1. From this figure, we see that the expected infection rate varies significantly across space types. In Office space, for example, the occupant density is quite low, as is the expected quanta generation rate; thus even at the minimum EOA rate of 1 ACH, the expected number of transmissions is only 0.5, and so the infector would need to be present for two days (16 h total) before they are expected to transmit their disease. By contrast, the high occupant density Restaurant has a much higher number of transmissions at 30 despite its higher minimum ventilation rate. Thus, in such spaces, additional infection control measures may be needed to achieve adequate infection risk. For the intermediate Classroom and Retail cases, number of transmissions is somewhat high but could be mitigated by requiring occupants to wear masks. In all cases, we can observe strong diminishing returns of additional EOA due to the inverse dependence.

**Table 1:** Parameters for space types. Exhaled quanta concentrations assume minimal talking for Office, some talking for Classroom or Retail, and loud talking for Restaurant (Bazant and Bush, 2021). Occupant densities and minimum ventilation rates are taken from ASHRAE Standard 62.1-2019 (ASHRAE, 2019).

| Space Type | Exhaled<br>Quanta<br>Conc.<br>(quanta/m<sup>3</sup>) | Occupant<br>Density<br>(#/1000 ft<sup>2</sup>) | Minimum<br>Ventilation<br>Rate<br>(ACH) |
| --- | --- | --- | --- |
| Office | 10 | 5 | 0.43 |
| Classroom | 25 | 35 | 2.35 |
| Retail | 25 | 15 | 1.16 |
| Restaurant | 100 | 70 | 3.52 |

**Figure 1:**
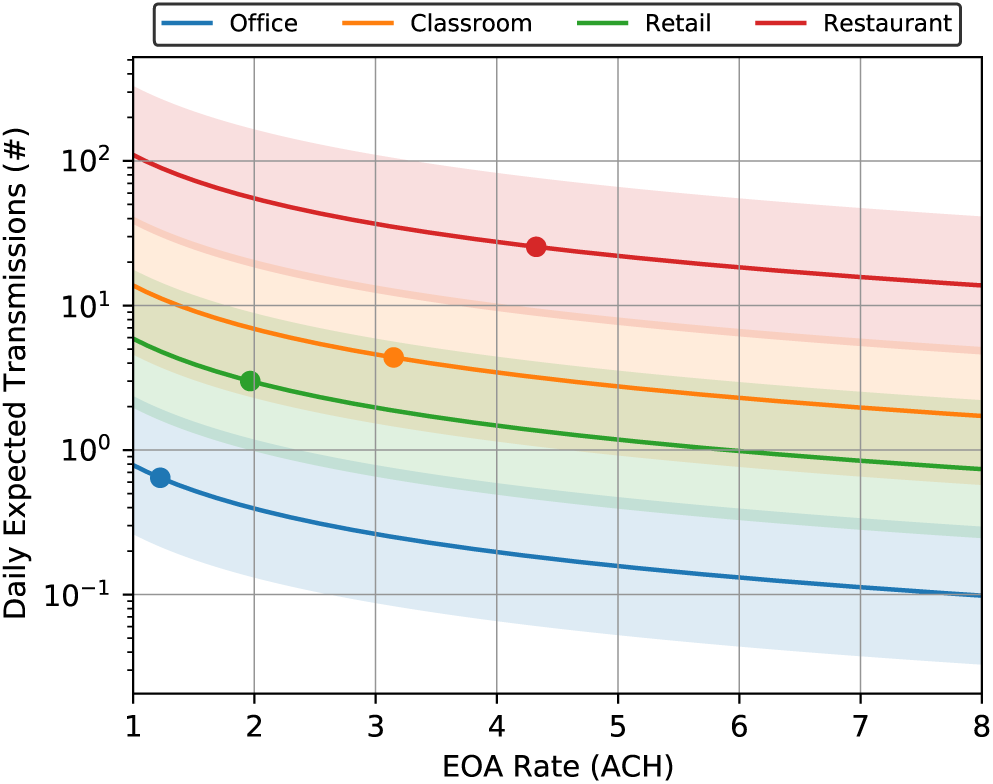
Number of transmissions versus EOA delivery for various space types. Indicated points on each curve show the minimum EOA rate expected for each space, which includes the ASHRAE-mandated minimum ventilation plus an additional 0.8 ACH corresponding to the passive physical processes. Shaded regions show a factor of 3 uncertainty in the exhaled quanta concentration.

As a second example case, we look at how the number of transmissions varies with outdoor air fraction and in-duct filter type. For this purpose, we take the Classroom case from the previous example and assume a total supply airflow of 6 ACH which would be typical for design conditions in the cooling season. Filtration efficiencies for each MERV size bin and averaged values (calculated via Eq. (2)) are shown in Table 2. To consider the effect of uncertainty, we assume each filter’s efficiency can vary between its minimum and maximum bin values from that table. The resulting EOA and transmission numbers are shown in Fig. 2. Most striking in this plot is the fact that increasing the outdoor air fraction is really only useful for low-efficiency filters. For the MERV 4 filter (with *η*_filter_ = 0.17) moving from minimum ventilation to 100% outdoor air increases the EOA rate from 4 to 6 ACH, which in turn reduces the infectious dose by about one third. By contrast, the highest-efficiency MERV 15 filter (with *η*_filter_ = 0.96), increasing the outdoor air fraction has almost no effect on EOA delivery or infection rate because almost all of the infectious particles are removed from the supply air stream regardless of ventilation. If weather is particularly warm, then increased ventilation is likely to require significant additional cooling energy (and may lead to discomfort in the space due to equipment capacity limitations). Thus, if this space has already been upgraded with a high-efficiency filter, then there is often little motivation to increase the ventilation rate above the minimum, unless *total* flow is also adjusted.

**Table 2:** Bin and average filtration efficiencies for MERV filter types. (ASHRAE Epidemic Task Force, 2021a).

| Filter Type | $\eta_{E1}$ (%) | $\eta_{E2}$ (%) | $\eta_{E3}$ (%) | $\eta_{\text{filter}}$ (%) |
| --- | --- | --- | --- | --- |
| MERV4 | 10.3 | 29.9 | 11.9 | 16.8 |
| MERV8 | 15.1 | 51.6 | 73.7 | 55.6 |
| MERV13 | 66.3 | 92.4 | 97.8 | 89.9 |
| MERV15 | 86.4 | 97.8 | 99.1 | 96.2 |

**Figure 2:**
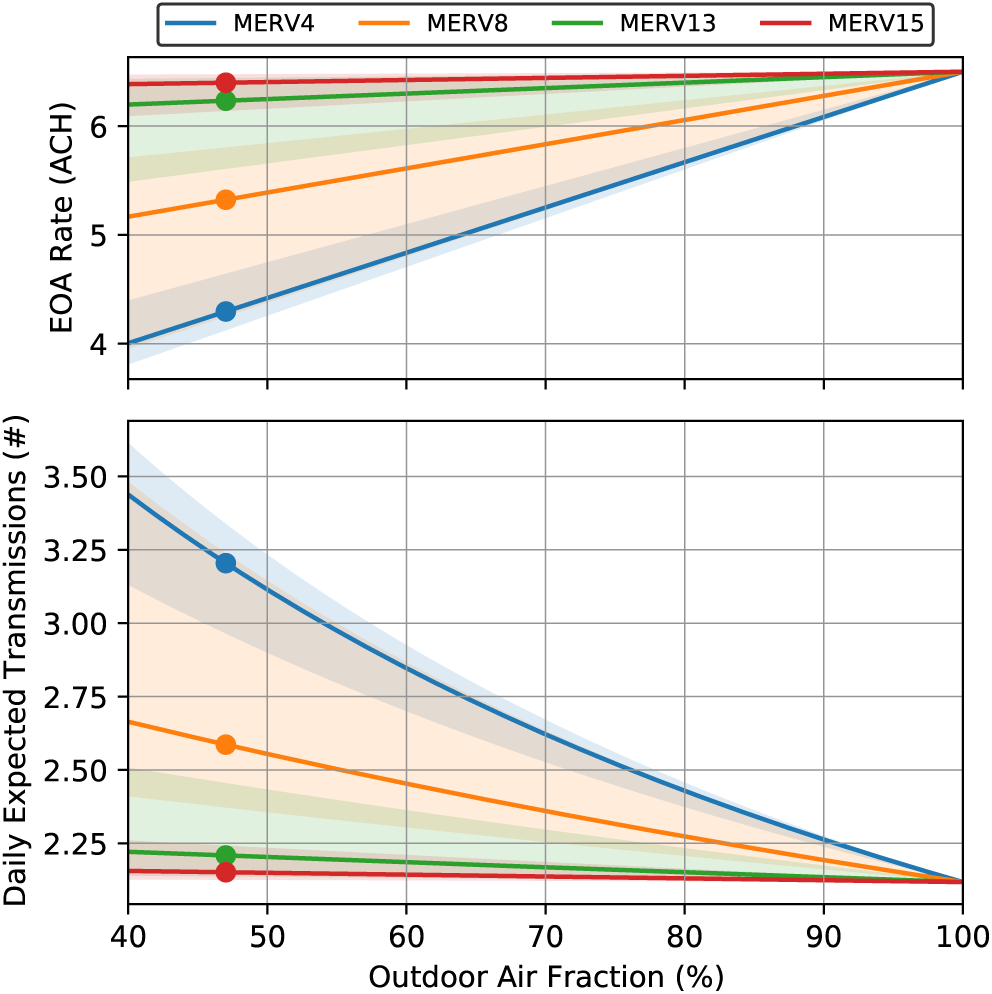
EOA delivery and number of transmissions versus outdoor air fraction for various filter types. Results assume 5 ACH total supply flow for the Classroom space type with 1.5 ACH of EOA for the passive physical processes. Shaded regions show uncertainty associated with actual filter efficiency being between the minimum and maximum bin values from Table 2.

## 3.2. Tradeoff Curve

We turn now to examine the tradeoff between energy consumption and transmission rate. As discussed in Section 2.5, two HVAC variables that can be adjusted to provide additional EAO are the supply temperature setpoint and the ventilation rate. Thus, to evaluate the operational landscape, we consider a grid over these two values, with the supply temperature setpoint ranging from 55 to 63 °F and the ventilation rate ranging from the ASHRAE minimum to two times that value (with the upper limit corresponding to nearly 100% outdoor air operation in this system). For each point on that grid, we evaluate the number of transmissions and energy consumption via the models in Section 2. Note that we treat the *ventilation rate* rather than the outdoor-air fraction as the independent variable so as to more clearly decouple from the effect of the supply temperature setpoint, but the most appropriate variable for a given system will depend on the specific outdoor-air control strategy being employed. Occupant density and quanta generation rate are taken from the Office case in the previous section, and the in-duct filter is rated MERV 8 (which provides some room for ventilation to reduce infection risk). Baseline zone and supply conditions are taken from an EnergyPlus simulation for the “Large Office” reference building (US Department of Energy, 2020). To account for time variation in indoor and outdoor conditions, we apply the calculations from Section 2 separately for each hour and then aggregate over the full 24 hour period (although the HVAC system is only active during the occupied period from 6am to 10pm). The results of these calculations are shown in Fig. 3.

**Figure 3:**
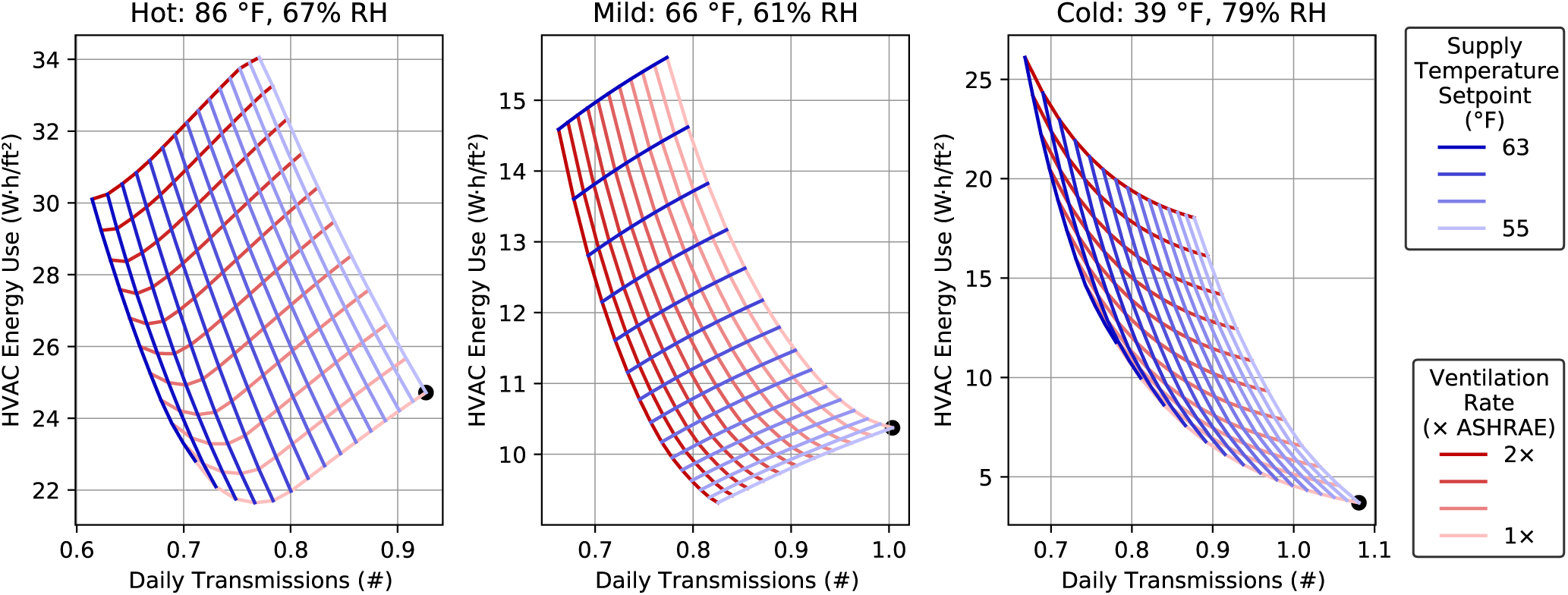
Example tradeoff curves between energy cost and transmission number for different weather conditions. Values are calculated based on 24 hours of operation for an office, with indicated weather giving average conditions over that span. Each red line holds the ventilation rate constant and varies supply temperature setpoint, while each blue line holds supply temperature setpoint constant and varies ventilation rate. Black points show baseline settings (with both variables at their minimum bounds).

In the Hot weather conditions, we see that the most efficient means of infection risk reduction is to increase the supply temperature setpoint while remaining at the lowest ventilation rate (the curve along the bottom of the region is red, which corresponds to varying the supply temperature setpoint). Intuitively this result makes sense, as when the ventilation rate is increased, additional energy must be consumed to condition the hot outdoor air. In this case, the modeling indicates an initial reduction in energy consumption as supply temperature is increased. This corresponds to the region where the reduction in latent load due to the higher supply temperature more than makes up for the increase in fan power needed deliver the additional flow. Once supply temperature is near the upper bound, additional infection mitigation requires increased ventilation, although by that point most of the available benefits have already been captured. By contrast, Mild conditions result in ventilation being the more efficient source of infection risk reduction: since the outdoor air is much closer to the supply temperature setpoint, little additional cooling energy is required (and indeed, energy consumption actually decreases slightly as the outdoor air fraction is reduced at low ventilation rates). Finally, under Cold weather the trends are qualitatively similar to Hot weather, except that now the increased energy consumption is to *heat* the extra outdoor air rather than cool it. Thus, adjusting the supply temperature setpoint is once again the more efficient variable to reduce infection risk. It should also be noted that in this case, the relative change in energy consumption is much higher because baseline energy consumption is much lower in Cold weather compared to Hot. We do note that the installation of energy-recovery ventilation equipment could significantly reduce the energy penalty associated with increased ventilation (MacNaughton et al., 2015), but of course such design changes would generally not be possible as a short-term infection control strategy.

Overall, this analysis illustrates that when considering HVAC measures to reduce infection risk, the current outdoor conditions play a significant role in determining the optimal strategy. Thus, buildings in milder climates may be able to rely on increased ventilation, whereas buildings in extreme climates should favor improved filtration and increased recirculation instead. In addition, building managers should reevaluate their chosen strategies, at least on a seasonal basis, to ensure that their buildings are not consuming unnecessary energy.

## 3.3. Space-by-Space Analysis

As a final example, we show how the infection risk and energy analysis can be used to compare different spaces within a building and prioritize supplemental disinfection. For this purpose, we consider a school building divided into six zones. Building dimensions and baseline HVAC variables are taken from an EnergyPlus simulation for the “Primary School” reference building (US Department of Energy, 2020). Key parameters for each zone are shown in Table 3. Note that the “Miscellaneous” zone includes administrative offices, a computer lab, and a library, while the three “Classrooms” zones each contain multiple classrooms served by the same AHU. As in the previous example, we account for time variation by performing analysis on an hourly basis and aggregating. The time-varying occupancy profile for each zone is shown in Fig. 4.

**Table 3:** Parameters for zones within the school building. Quanta concentrations are chosen based on expected activity and vocalization levels for each zone.

| Zone | Max<br>Occupancy<br>(#) | Exhaled<br>Quanta Conc.<br>(quanta/m<sup>3</sup>) | Area<br>(1000 ft<sup>2</sup>) | Volume<br>(1000 ft<sup>3</sup>) | Mean<br>Supply Flow<br>(1000 cfm) | Mean<br>Ventilation<br>Fraction (%) |
| --- | --- | --- | --- | --- | --- | --- |
| Classrooms 1 | 249 | 25 | 15.5 | 203.8 | 6.14 | 86% |
| Classrooms 2 | 212 | 25 | 13.4 | 175.9 | 4.93 | 88% |
| Classrooms 3 | 200 | 25 | 12.7 | 167.0 | 4.89 | 86% |
| Gym | 102 | 100 | 3.8 | 50.4 | 2.57 | 88% |
| Cafeteria | 216 | 100 | 3.4 | 44.5 | 4.80 | 100% |
| Miscellaneous | 182 | 10 | 23.3 | 305.3 | 5.68 | 83% |

**Figure 4:**
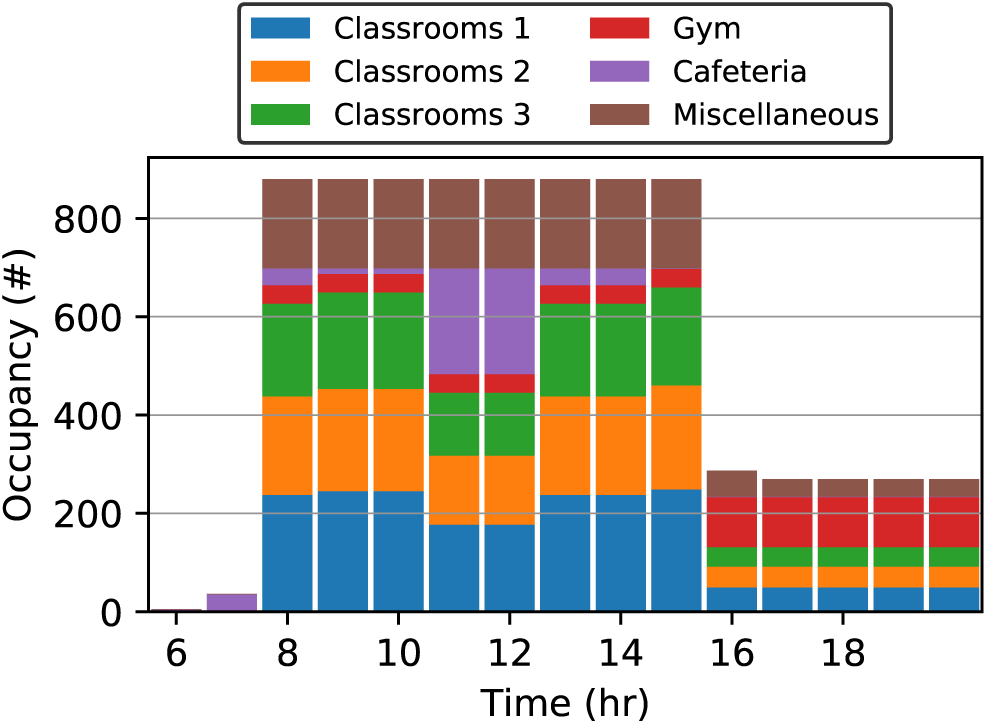
Time-varying occupancy by zone for the school building. Occupancy is zero outside of the indicated time range.

We assume for this example that the school has decided to install eleven portable filtration devices with HEPA filters, each providing 1,000 CFM of supplemental EOA to the zones in which they are located. (Such devices have a larger capacity than would be expected in low-occupancy or residential settings but are more typical of high-occupancy or commercial applications.) The purpose of this analysis is to identify the zones in which each device should be installed in order to maximize effectiveness. To analyze where in-zone filtration would be most helpful, we start by assuming that 1% of occupants in the building are actively infectious. By fixing the fraction of infectors, the average number of infectors in each zone is equal to 1% of that zone’s time-varying occupancy, and thus we inherently account for variations in occupant density. We can then identify the optimal zone for each disinfection device by choosing the zone where that device would lead to the biggest reduction in total transmissions *N*_*T*_ for the building. In this context, it is important *not* to compare spaces by their reproductive number *R*, as a specific zone could have a very high value of *R* but pose little overall transmission risk due to having a very small number of occupants. Because we consider the zones to be isolated from one another, we can assess each zone individually under baseline conditions and then with varying levels of supplemental *f* ^EOA^ from the new filtration units. We then calculate the reduction in transmissions as each filtration unit is added. The results of these calculations are shown in Fig. 5.

**Figure 5:**
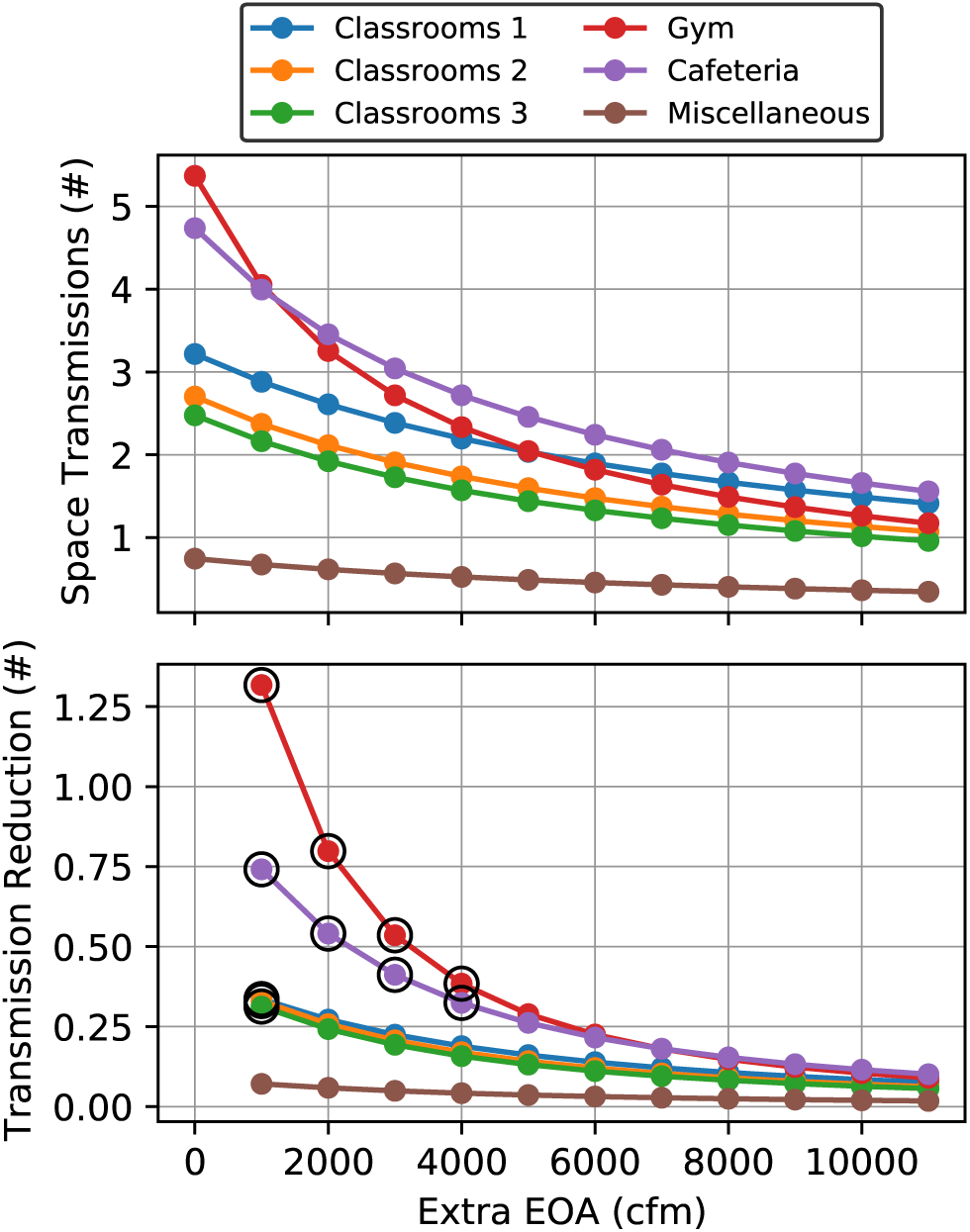
Total transmissions and reduction in transmissions for up to eleven in-zone filtration units in each zone of the school. Circled points indicate the eleven locations that have the highest reduction, which are thus the optimal locations to place the devices.

From our calculations, we see that the highest reduction in transmission is achieved by placing four units in the Gym, four units in the Cafeteria, and one unit each in the three Classroom zones. This result is perhaps counterintuitive, as the building occupants spend the overwhelming majority of their time in the Classroom zones. However, due to the higher quanta emission rates, total infection risk is higher in the first two spaces. Combining that effect with space geometry thus makes these zones better targets for supplemental disinfection. For this example case, placing the devices in these locations reduces the total number of transmissions from 19.3 to 13.2, which is a 31% reduction from the baseline value.

Finally, we suppose that the school wants to estimate the energy impact of installing the additional devices and determine whether a similar level of infection risk reduction could be achieved via in-duct filtration. To this end, we calculate the energy consumption of each zone’s HVAC system with the additional in-zone filtration devices (each drawing 650 W of electricity while active) assuming the current MERV8 in-duct filter and alternatively an upgraded HEPA in-duct filter. We show the results of these calculations in Fig. 6. The main finding is that, although the induct HEPA filter requires only a small amount of extra electricity consumption (generally 2% to 4%), overall disease transmission risk is only slightly reduced. The main limitation is that, as shown in Table 3, the zones are already near 100% outdoor air in the current conditions, and thus the filters have very little “dirty” air left to filter. (The cafeteria in particular receives no benefit due to already operating at 100% outdoor air.) Thus, the in-zone filtration devices are the more effective way to provide disinfection for the school, although they do come with significantly increased energy consumption.

**Figure 6:**
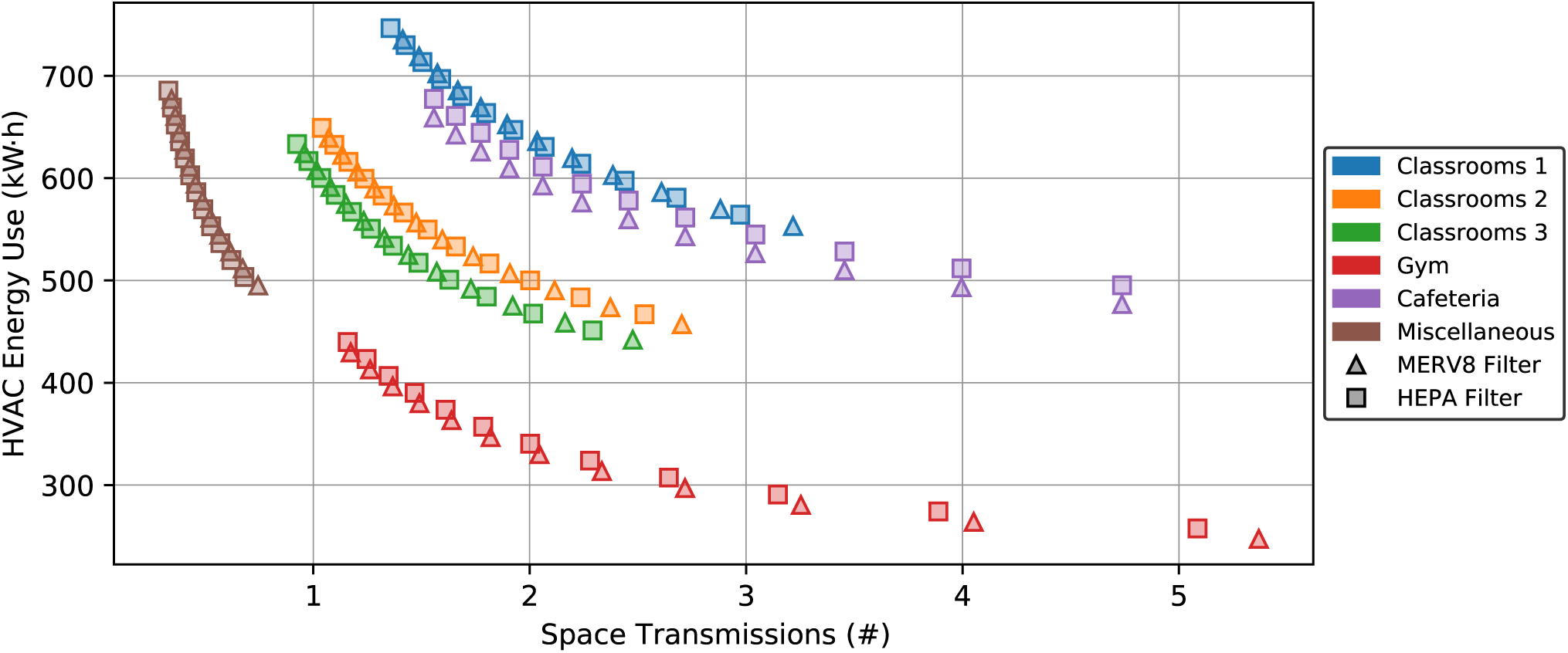
HVAC energy consumption and number of transmissions for each zone assuming up to eleven in-zone disinfection devices for two different in-duct filter types. The rightmost points for each zone correspond to no in-zone devices, while the leftmost points show the effects of all eleven devices.

The main point of this example is to illustrate that practical analysis of infection risk and energy consumption is relatively straightforward despite the many factors that must be considered. In particular, a thorough analysis of infection risk must account for time, occupancy, and activity level in each space to ensure a common basis of comparison, but the key parameters are often readily obtainable. More detailed analysis could also consider changes to the operation of the HVAC system to provide increased airflow to the space, which could be a more energy-efficient source of disinfection depending on weather and other conditions.

## 4. Conclusions and Future Directions

In this paper, we have presented simple models to quantify the risk of airborne disease transmission in buildings alongside the energy costs of various HVAC actions that can help mitigate that risk. By modeling a hypothetical concentration of infectious particles in the air, the expected infection rate in a space can be determined from a small number of parameters, including occupancy level, occupant behavior, and HVAC system operation. By formulating all disinfection mechanisms using the common basis of equivalent outdoor air, the incorporation of new ventilation, filtration, or other clean-air sources is straightforward. By accounting for both infection risk *and* energy consumption associated with various operational decisions (e.g., changing supply temperature, ventilation rate, or filter type), these models allow building managers to make informed decisions about which strategies to employ consistent with current priorities. We have illustrated through examples that the proposed analysis can yield nontrivial insights and that the best course of action can vary significantly from building to building and even within the same building depending on weather conditions or occupant behavior.

In the future, we hope to extend this analysis to focus on more general aspects of indoor air quality rather than just airborne disease transmission. Indoor contaminants like CO_2_, VOCs, and particulates behave similarly (though not identically) to the infectious particles modeled in this paper, and thus the framework can be extended accordingly. Where direct measurements of indoor contaminant concentrations are available, the indoor generation rates for each species can be estimated directly from data rather than having to estimate based on occupant behavior (as is the case for the exhaled quanta concentration in this work). The overall goal would be to provide quantitative guidance to building managers to help assess the tradeoff between occupant wellness and energy consumption. Indeed, previous studies have shown reducing indoor CO_2_ concentrations (and improving IAQ in general) can provide significant health and cognition benefits to building occupants (MacNaughton et al., 2015), but the additional energy consumption associated with increased ventilation can be very high depending on climate. Given this conflict between energy sustainability and occupant wellness, it is thus critical to facilitate informed decision-making in this area, just as in the airborne transmission case considered in this work.

## Data Availability

All data is included in the manuscript or referenced works.

